# MRI-Based Pressure Gradient Mapping in Patient-Specific Models of Coarctation of the Aorta

**DOI:** 10.64898/2026.05.27.26353898

**Authors:** Priya J. Nair, Lorenzo Ferrari, Michael Loecher, Charles McGrath, Carlos A. Castillo Passi, Alison L. Marsden, Daniel B. Ennis

**Affiliations:** Department of Bioengineering, Stanford University, Stanford, CA, USA; Stanford Cardiovascular Institute, Stanford University, Stanford, CA, USA; Department of Pediatrics - Cardiology, Stanford University, Stanford, CA, USA; Maternal and Child Health Research Institute, Stanford University, Stanford, CA, USA; Institute for Computational and Mathematical Engineering, Stanford University, Stanford, CA, USA; Department of Cardiothoracic Surgery, Stanford University, Stanford, CA, USA; Department of Radiology, Stanford University, Stanford, CA, USA; Division of Radiology, VA Palo Alto Healthcare System, Palo Alto, CA, USA

**Keywords:** aortic coarctation, hemodynamics, 4D-Flow MRI, computational fluid dynamics

## Abstract

**Purpose:** Accurate assessment of the pressure gradient (**Δ*P***) across aortic coarctation (CoA) is critical for determining disease severity and the need for intervention. Current non-invasive methods are unreliable, while invasive catheterization remains the clinical gold standard. This study evaluates a novel MRI acquisition strategy, 4D-FlowP, that simultaneously encodes blood velocity and acceleration to enable reliable non-invasive pressure gradient mapping in CoA.

**Methods:** Patient-specific compliant aortic phantoms were created from clinical MRI data of two patients with CoA. Additional geometries were synthetically generated by increasing stenosis severity. Phantoms were studied in an MRI-compatible flow loop under physiologically realistic flow and pressure conditions. Pressure gradients were estimated using conventional 4D-Flow MRI, 4D-FlowP, and fluid–structure interaction (FSI) simulations. Results were compared against ground-truth catheter-based measurements across multiple flow rates and stenosis severities.

**Results:** Conventional 4D-Flow consistently underestimated **Δ*P*** (slope = 0.63, ***R***^**2**^=0.75) relative to catheter measurements. In contrast, 4D-FlowP demonstrated substantially improved agreement (slope = 0.95, ***R***^**2**^=0.75). FSI simulations showed the highest overall agreement with catheter-derived **Δ*P*** (slope = 1.14, ***R***^**2**^=0.82). Scan times for 4D-FlowP were comparable to 4D-Flow (26 vs. 24 minutes).

**Conclusion:** 4D-FlowP enables a more accurate MRI-based pressure gradient mapping in CoA than conventional 4D-Flow, when compared to ground truth catheter measurements. These findings support further *in vivo* evaluation of 4D-FlowP as a non-invasive alternative for functional assessment of CoA severity.

## 1 Introduction

Coarctation of the aorta (CoA) is a form of congenital heart disease characterized by a constriction of the aorta, typically just distal to the origin of the left subclavian artery. CoA accounts for 6-8% of all congenital heart diseases [1], with an incidence of approximately 3 cases per 10,000 births [2]. The vessel narrowing increases resistance to blood flow, leading to hypertension proximal to the CoA, and an elevated pressure gradient (Δ*P*) across the CoA. The increased resistance to flow also results in higher ventricular afterload. If left untreated, CoA can lead to premature coronary artery disease, ventricular dysfunction or failure, aortic aneurysms, aortic dissection, and increased risk of cerebral aneurysms and stroke [3].

For pre-operative evaluation of CoA severity, magnetic resonance imaging (MRI) or computed tomography (CT) are often used for anatomical assessment, which allows for geometric assessment of CoA, but these do not measure the functional consequences. Functional assessment is performed by measuring Δ*P* across the CoA. The current clinical indication of a severe CoA warranting corrective intervention is Δ*P ≥* 20 mmHg [4]. The gold-standard method for measuring Δ*P* is invasive cardiac catheterization.

Non-invasive clinical methods for estimating Δ*P* include Doppler echocardiography and, historically, estimating a difference in cuff blood pressure (BP) measurements between the arms and legs. These non-invasive methods, however, are known to be less accurate than catheterization. Doppler echocardiography, with both the simplified and modified Bernoulli’s equation, overestimates Δ*P* by 41% on average [5]. The difference in BP between the upper and lower extremities is also reportedly unreliable compared to the gold-standard, with one study reporting an average error of 72% in the Δ*P* estimated from cuff BP versus cardiac catheterization. [6]. Therefore, accurate clinical assessment of Δ*P* to determine CoA severity currently requires an invasive and costly catheter-based diagnostic test. Improved non-invasive methods for functional assessment of CoA severity offer the potential to avoid drawbacks and risks of invasive catheterization (including bleeding, infection, and exposing patients to radiation and contrast agents) and also lower the cost of patient care.

The use of computational fluid dynamics (CFD) to assess hemodynamics in patients with CoA has been explored in the past [7–12], but these methods can be time-consuming and therefore not always clinically applicable. The ideal method for Δ*P* estimation would be one that could be seamlessly integrated into the typical clinical workflow.

In principle, phase contrast MRI (PC-MRI) can measure *in vivo* pressure gradients [13, 14]. Generally, this is done with velocity encoding techniques such as 4D-Flow. Given a measured velocity field 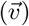, the pressure gradient Δ*P* can be computed by evaluating the Navier-Stokes equation for conservation of momentum:

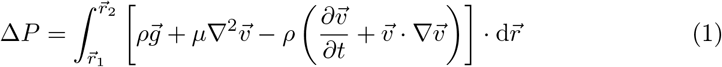

Therein, *ρ* is the density of the fluid, 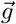 is the gravitational acceleration, *µ* is the dynamic viscosity of the fluid, 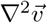 is the Laplacian of the velocity field (viscous diffusion term), 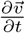 is the temporal acceleration, and 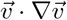 is the convective acceleration. The variables 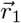 and 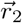 represent arbitrary spatial coordinates in the fluid domain. While this technique of estimating Δ*P* using MRI has shown good accuracy [13, 14], it is highly sensitive to noise in the velocity data due to its reliance on spatial and temporal gradients. Previous work showed that by directly encoding acceleration instead of velocity, noise sensitivity was decreased, and therefore the accuracy of pressure gradient mapping can be improved [15].

In this work, we aim to demonstrate the utility of a novel numerically optimal method (“4D-FlowP”) for simultaneously encoding velocity and acceleration in an MRI acquisition for improved pressure gradient mapping. We validate the accuracy of Δ*P* measurements determined using 4D-FlowP by comparing to estimates obtained from 4D-Flow and ground-truth catheter-based measurements in a set of patient-specific phantoms with CoA over a range of flow rates. Additionally, we compare our *in vitro* measurements with results from corresponding fluid-structure interaction (FSI) simulations for further validation.

## 2 Materials and Methods

In this section, we summarize the methods used to create patient-specific compliant CoA phantoms and the protocol used for MRI data acquisition. We also provide background on the 4D-FlowP technique and the methods used to perform FSI simulations. Finally, we describe the analysis performed to compare Δ*P* estimates derived from 4D-Flow, 4D-FlowP, and FSI simulations to those acquired by direct measurement through catheterization.

### 2.1 Patient Data Acquisition

Under a protocol approved by the Stanford Institutional Review Board, patients with CoA were retrospectively identified from the Lucile Packard Children’s Hospital at Stanford. Patients with native and recurrent CoA were included. Inclusion criteria for the cohort required an imaging exam (with 4D-Flow MRI) and invasive pressure measurements acquired via cardiac catheterization. While previous studies have explored the impact of bicuspid aortic valves and collateral formation on the hemodynamics in patients with CoA [16, 17], patients with aortic valve stenosis and/or extensive collaterals were excluded from this study. We obtained retrospective 4D-Flow MRI datasets for two patients with CoA. Invasive measurements of pressure via cardiac catheterization in the ascending aorta (AAo) and descending aorta (DAo) were obtained, in addition to cuff BP and heart rate. Informed consent was not required for this retrospective clinical data collection.

### 2.2 3D Model Construction and Phantom Generation

MRI magnitude images were imported into SimVascular (simvascular.org) [18]. Centerlines were manually drawn through the aorta and the branches arising from the aortic arch: the brachiocephalic trunk (BCT), the left common carotid artery (LCA), and the left subclavian artery (LSA). 2D contours were manually drawn along the centerlines to trace the blood vessel lumen and then lofted to produce a patient-specific model of the blood volume (Figure 1A). To evaluate the accuracy of 4D-FlowP in a wider range of Δ*P*, the severity of the stenosis was increased for each of the patients using svMorph [19], resulting in a total of four 3D geometries (Figure 1B).

**Fig. 1.**
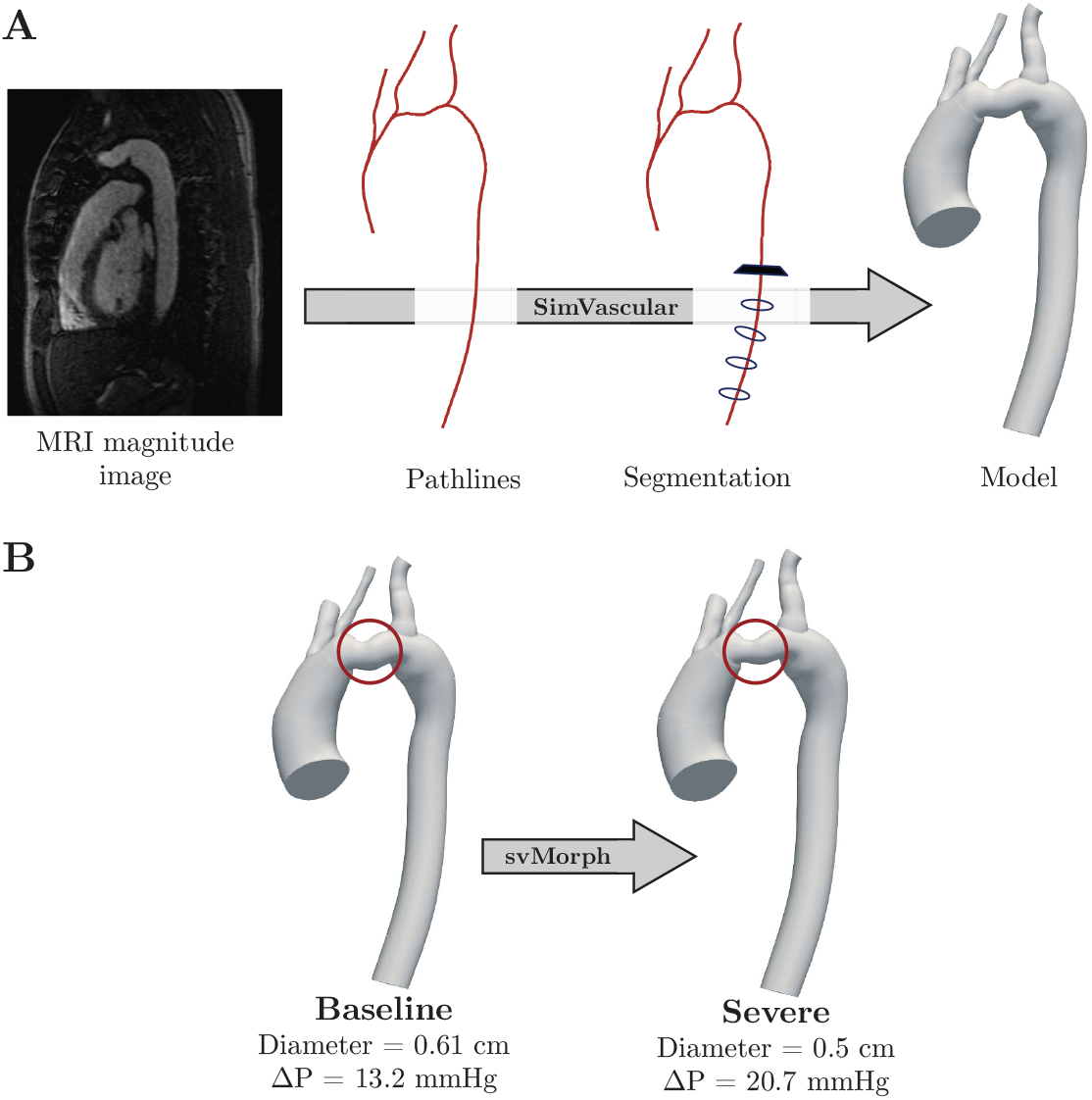
(A) Model generation pipeline in SimVascular. (B) Use of the svMorph tool to increase the severity of the stenosis in the 3D model.

Meshmixer (Autodesk) was used to generate the vessel wall (wall thickness = 2.5 mm) and to add cylindrical caps (length = 20 mm) at the inlets and outlets to enable connection to custom barbed transition elements that connect the aortic phantom to tubing. A photopolymerization 3D printer (J735 PolyJet, Stratasys) with a compliant printing material (Agilus30, Stratasys) was used to manufacture phantoms with compliant walls from the patient-specific aorta geometries (Figure 2) with an elastic modulus approximating the human aorta (*E* = 1.2 MPa) [20, 21]. The printed phantoms were coated with a thin conformal coating (DOWSIL 3-1953) to prevent fluid absorption.

**Fig. 2.**
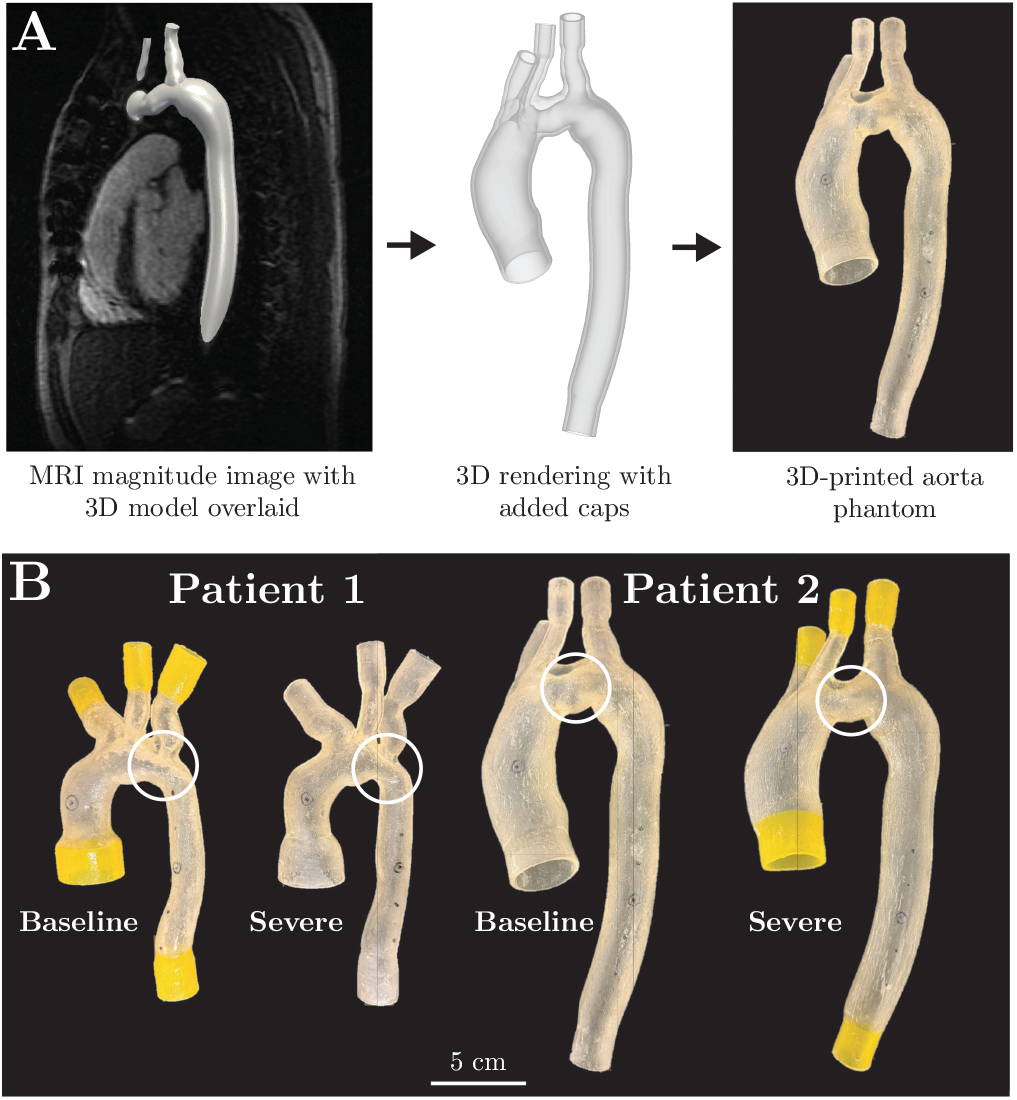
(A) Patient-specific phantom generation process. (B)3D-printed patient-specific phantoms of CoA with compliant walls. White circles indicate location of CoA

### 2.3 *In Vitro* Cardiovascular System Emulator

We used our cardiovascular system emulator (Figure 3) to enable *in vitro* 4D-Flow imaging under patient-specific physiological flow and pressure conditions. Custom 3D-printed barbed connectors with tapered transitions were used to connect the inlets and outlets of the 3D-printed phantom to tubing. The phantom was embedded in a gel block (ClearBallistics) that provided a fixed positioning reference and served as static “tissue” for background phase correction. The working fluid was a glycerol-water mixture (ratio = 40/60) with density (1.1 *g/cm*^3^) and dynamic viscosity (0.0042 Pa · s), closely matching that of human blood. A *T*_1_-shortening contrast agent (ferumoxytol, concentration = 0.75 mL/L) was added for increased signal-to-noise ratio (SNR). The phantom inlet was connected to the outflow of the pump unit (CardioFlow 5000 MR, Shelley Medical Technologies). The outlets emptied into a reservoir that then connected to the pump inflow resulting in a closed flow circuit.

**Fig. 3.**
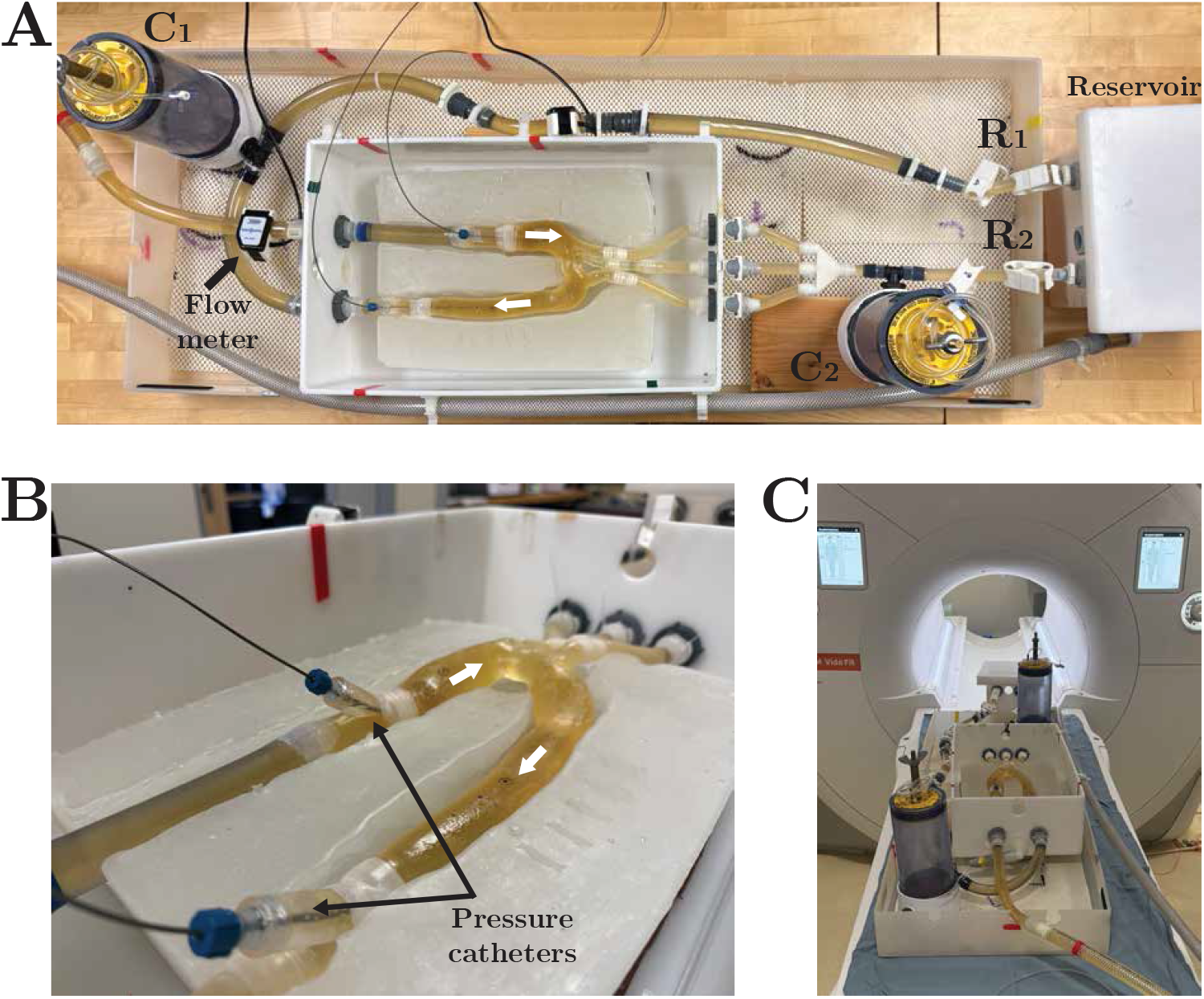
(A) MRI-compatible *in vitro* cardiovascular system emulator with an embedded 3D-printed compliant patient-specific CoA model instrumented for direct pressure and flow rate measurements. The system includes tunable fluid capacitors (*C*_1_ and *C*_2_) and resistors (*R*_1_ and *R*_2_) to match patient-specific hemodynamics. White arrows indicate the direction of flow. (B) Zoomed in view of the model-specific gel block with embedded aortic phantom and pressure catheter ports (blue) at the inlet and outlet. (C) The MRI-compatible cardiovascular system emulator prepared for a 3T MRI (Vida Fit, Siemens) study on the scanner bed.

The inflow waveform (averaged over the vessel cross-sectional area, cm^2^) was derived from the patient’s *in vivo* 4D-Flow MRI exam. The original waveform was measured by manually drawing a contour of the inlet (just past the aortic sinus) on background-phase corrected 4D-Flow images acquired using a vendor sequence. This was used to quantify the time-resolved flowrate (mL/s) averaged over the cross-sectional area. The original flowrate waveform was spline-interpolated and down-scaled to meet the pump’s instantaneous flow rate limit of 300 mL/s (*Q*_2_), and discretized (Δ*t* = 10 ms) over a cardiac cycle length matching the patient’s clinical heart rate (HR) measurement. The applied flowrate waveform was then measured using two clamp-on ultrasonic flow probes (Transonic Systems Inc.), one placed upstream of the AAo inlet and one placed downstream of the DAo outlet. Pressures in the system were manually adjusted using resistive and capacitive elements to match pressure measurements derived from *in vivo* catheterization. Mean pressure in the system was adjusted using clamp-on pinch valve resistances at the outlets (*R*_1_ and *R*_2_). The ratio of *R*_1_ to *R*_2_ was also adjusted to match flow volume (mL/beat) splits across the the model outlets to measurements derived from *in vivo* 4D-Flow MRI.

Pulse pressure was controlled using capacitance elements (*C*_1_ and *C*_2_). The capaci-tors were designed as cylindrical towers with sealed air compression chambers in which the enclosed air volume (height = 16 cm, diameter = 10.2 cm) dictated the amount of downstream capacitance. An additional reduced flow rate (*Q*_1_ = 0.5*Q*_2_) was also studied. Higher flow rates were not possible due to pump limitations. *R*_1_, *R*_2_, *C*_1_, and *C*_2_, once tuned for the baseline geometry, were kept constant for each patient even with increasing stenosis severity or varying cardiac output. Pressure transducers (MicroTip SPR-350S, Millar) were inserted through Tuohy-Borst adapters at the AAo inlet and DAo outlet. After priming and tuning the system, ground truth pressure measurements were acquired in the AAo proximal to the origin of the BCT and in the DAo two vertebral spaces above the diaphragm to match clinical measurement locations in pre-defined slices based on clinical imaging. Markers were added to the gel to indicate the slice locations at which catheter measurements were acquired such that they were visible in acquired MR images. Each pressure signal was received at the data acquisition (DAQ) system (PowerLab, AD Instruments) through a bridge amplifier front-end (FE224 Quad Bridge, ADInstruments) and recorded using LabChart software (ADInstruments). Δ*P*_Cath_ was calculated as the maximum difference in pressure between the AAo and DAo averaged over five cardiac cycles.

### 2.4 4D-FlowP

The time varying magnetic field gradients in MRI cause the spins (hydrogen nuclei) to accumulate phase. The phase (*ϕ*) that a moving spin accumulates depends on: 1) the spin’s position 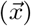, velocity 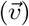, and acceleration 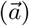, plus 2) the zeroth 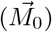, first 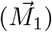, and second 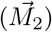 moments of the gradient waveform (higher order terms are ignored):

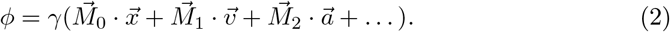

Here, the n^th^ moment of the gradient is defined as

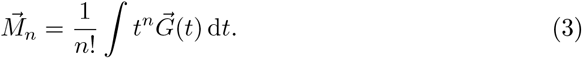

In a standard anatomy scan, the gradients are designed such that 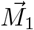 and 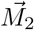 are zero, so the phase encodes the position to form an image. In PC-MRI, 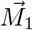 is designed to be non-zero so the phase encodes the velocity. In 4D-FlowP, both 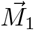 and 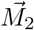 are designed to be simultaneously non-zero such that the phase encodes both the velocity and acceleration. Gradient waveforms were designed with an optimization framework (GrOpt)[22], which enabled the design of time optimal encoding gradients. The result is a set of unique and non-collinear 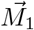 and 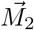 while staying within the system hardware limits. 4D-FlowP uses seven encoding directions (including an off-resonance phase measurement, *ϕ*_off_ with no encoding). We then solve the system in Eqn. 4 to calculate velocity and acceleration components along the *x, y*, and *z* directions:

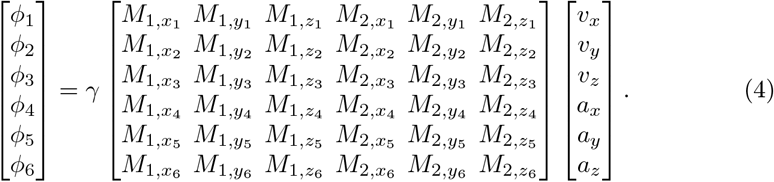

Here, *ϕ*_1_ to *ϕ*_6_ refer to measured phases after correction by *ϕ*_off_ ; and 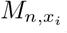 refers to the n*th* gradient moment along the i*th* direction. ∥ *M*_1_ ∥ and ∥*M*_2_ ∥ per encoding were selected based off of the prescribed *v*_*enc*_ and *a*_*enc*_ respectively. The directions of each encode were optimized to minimize the condition number and the encoding time for the gradient waveforms.

### 2.5 MRI Data Acquisition

We performed our imaging experiments using a 3T MRI scanner (Vida Fit, Siemens Healthineers) with a 32-channel spine coil and an 18-channel chest coil. We acquired: (1) high-resolution 3D spoiled gradient echo (SPGR) anatomical images, (2) 2D-PC flow measurements at pre-defined landmark slices (under pulsatile flow), and (3) 4D-Flow and 4D-FlowP flow and relative pressure measurements (under pulsatile flow). The pump trigger signal was used for retrospective cardiac gating (direct input to scanner).

#### 2.5.1 2D-PC MRI

Two-dimensional (2D) imaging planes perpendicular to the flow direction (and vessel centerline) were defined at three locations: (1) ascending aorta inlet, (2) descending aorta outlet, and (3) across the three branching head and neck vessels (BCT, LCA, and LSA). Images were acquired with the following scan parameters: matrix size 192×144, spatial resolution 1.875 mm × 1.875 mm × 6 mm, flip angle 20°, TE 2.5 ms, TR 6.01 ms, and *v*_*enc*_ 150-350 cm/s, BW 449 Hz/pixel, 3 segments, GRAPPA R=2, Cartesian, temporal resolution 37.5 ms, scan time 12-14 seconds.

The flow waveform measured using 2D-PC subsequently defined the inflow waveform in the FSI simulations described in Section 2.6.

#### 2.5.2 4D-Flow MRI and 4D-FlowP

The 4D-Flow protocol used a fully sampled Cartesian 4D-Flow sequence with a velocity encoding (*v*_*enc*_) range of 150 - 300 cm/s for Patients 1 and 2. For each aortic geometry, the *v*_*enc*_ was chosen to maximize velocity-to-noise ratio during systole while avoiding phase-wrapping artifacts, i.e. just above peak systolic velocities as measured by preceding 2D-PC. For 4D-FlowP, we used the same *v*_*enc*_ as the corresponding 4D-Flow acquisition. An additional acceleration encoding (*a*_*enc*_) was also defined with a range of 200 - 400 m/s^2^ for Patients 1 and 2. The parameters used for acquiring these images are outlined in Table 1. The specific *v*_*enc*_ and *a*_*enc*_ values used for each experiment are provided in the Appendix (Table 3).

**Table 1.** Imaging acquisition parameters.

| Parameter | 4D-Flow | 4D-FlowP |
| --- | --- | --- |
| Matrix Size | 220 × 176 × 64 |  |
| FOV [mm<sup>3</sup>] | 396 × 316.8 × 108.8 |  |
| Spatial Resolution | 1.8 mm × 1.8 mm<br>× 1.8 mm | 1.7 mm × 1.8 mm<br>× 1.8 mm |
| Flip angle [°] | 8 |  |
| Temporal Resolution<br>[ms] | 60 |  |
| $v_{enc}$ [cm/s] | 150 - 300 | |
| $a_{enc}$ [m/s<sup>2</sup>] | – | 200 - 400 |
| TE [ms] | 2.55 - 2.85 | 4.98 - 5.50 |
| TR [ms] | 5.05 - 5.35 | 7.59 - 8.21 |
| Encode Directions | 3+1 | 6+1 |
| Bandwidth [Hz/pixel] | 490 | 568 |
| Undersampling [R] | 2 | 5.25 |
| Scan Time [min:sec] | 21:37 - 26:53 | 24:47 - 26:49 |

**Table 2.**
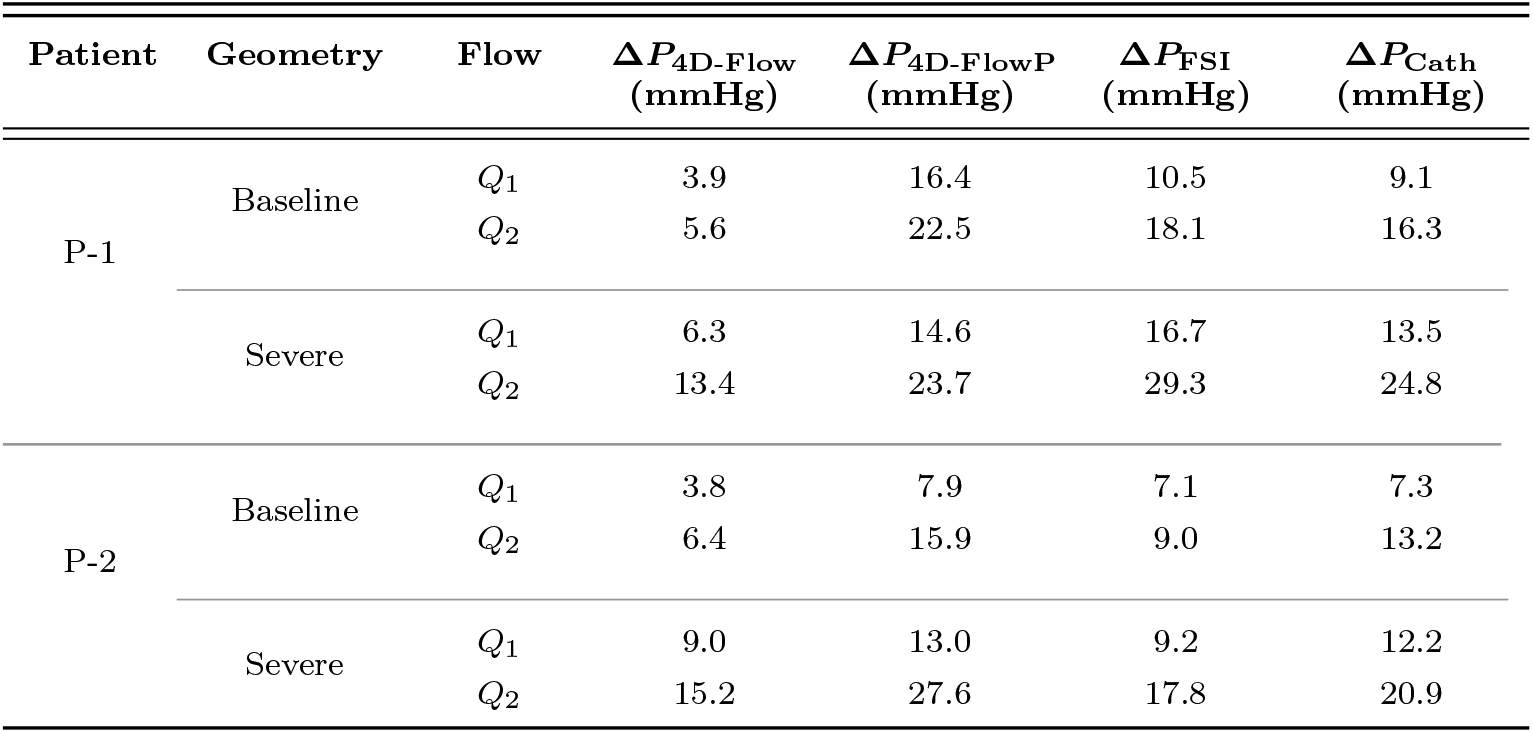
Peak Δ*P* between AAo and DAo in N=2 patients at two stenosis severities (Baseline and Severe) and two flow rates (*Q*_1_ and *Q*_2_) estimated from 4D-Flow, 4D-FlowP, and FSI, with catheter measurements shown for reference.

| Patient | Geometry | Flow | $\Delta P_{4D-Flow}$<br>(mmHg) | $\Delta P_{4D-FlowP}$<br>(mmHg) | $\Delta P_{FSI}$<br>(mmHg) | $\Delta P_{Cath}$<br>(mmHg) |
| --- | --- | --- | --- | --- | --- | --- |
| P-1 | Baseline | $Q_1$ | 3.9 | 16.4 | 10.5 | 9.1 |
| | | $Q_2$ | 5.6 | 22.5 | 18.1 | 16.3 |
| | Severe | $Q_1$ | 6.3 | 14.6 | 16.7 | 13.5 |
| | | $Q_2$ | 13.4 | 23.7 | 29.3 | 24.8 |
| P-2 | Baseline | $Q_1$ | 3.8 | 7.9 | 7.1 | 7.3 |
| | | $Q_2$ | 6.4 | 15.9 | 9.0 | 13.2 |
| | Severe | $Q_1$ | 9.0 | 13.0 | 9.2 | 12.2 |
| | | $Q_2$ | 15.2 | 27.6 | 17.8 | 20.9 |

Raw data was retrospectively binned to achieve a phase interval of 40ms and reconstructed with the *SenseRecon* functionality of SigPy [23, 24]. No additional spatial or temporal regularization was applied. Gradient distortion correction was then applied based on scanner specific parameters and background phase was corrected with a third order polynomial fit to the static gel. Finally, velocity and acceleration images were retrieved by applying the pseudo-inverse of the encoding matrix (Eqn. 4).

Pressure gradients were computed using the incompressible Navier-Stokes equation (Eqn. 1), with acceleration derived from velocity gradients (4D-Flow) or direct acceleration measurements (4D-FlowP). The gravitational term and viscous term in Eqn. 1 are considered negligible. To recover the scalar pressure field, the pressure gradient field was then used to formulate the pressure Poisson equation,

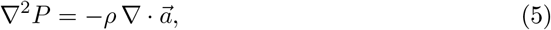

which was discretized over the segmented fluid domain defined by a binary mask. The resulting linear system was assembled using first order finite differences and solved using a sparse least-squares solver (Least Squares Minimal Residual, LSMR) [25]. Δ*P* _4D-Flow_ and Δ*P* _4D-FlowP_ were calculated as the maximum difference in pressure between the AAo and DAo measured using fully sampled 4D-Flow and 4D-FlowP respectively, averaged over the cross-sectional area at the same slice locations where ground-truth catheter measurements were acquired *in vitro*.

### 2.6 FSI Simulations

The same aortic geometries described in Section 2.2 were used. The vessel wall was modeled as a thin membrane. The 3D blood volume was meshed using tetrahedral elements. We also incorporated a boundary layer mesh consisting of three layers to resolve the high velocity gradient at the wall. In the region of interest (at the CoA and the region immediately distal to it), we further refined the mesh to capture the high-velocity jet created by the stenosis as it travels through the vessel narrowing and subsequent post-stenotic dilation. Based on a mesh convergence study, meshes with *∼*2 million linear tetrahedral elements were selected to ensure convergence of Δ*P* measurements.

We compared Δ*P* measurements acquired from *in vitro* catheter measurements and MRI against those from 3D FSI simulations. The inflow waveform measured using the 2D-PC slice at the ascending aorta inlet was prescribed as the inlet boundary condition of the FSI simulation. A three-element Windkessel model (proximal resistance *R*_p_, capacitance *C*, distal resistance *R*_d_) was imposed at each of the outlets in an open-loop fashion.

The total peripheral resistance (*R*_total_) was determined using mean inflow and mean aortic BP (*P*_mean_) measured *in vitro* as follows:

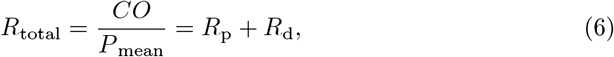

where the capacitance *C* and resistance ratio *R*_p_:*R*_d_ were iteratively tuned to match flow splits and pressures achieved on the bench within 5 mmHg using the approach outlined in Nair *et al*. [26]. The tuning was performed only for the baseline geometry; outlet boundary conditions were kept constant for the subsequent geometries when the stenosis severity was increased and for the varying inflow rates. We initialized all the 3D simulations with tuned 0D simulations, thereby allowing the 3D simulations to converge faster than they would have without any initialization [26, 27].

We performed 3D FSI simulations using the coupled momentum method (CMM) with svSolver, SimVascular’s finite element solver for fluid-structure interaction between an incompressible, Newtonian fluid and a linear elastic membrane for the vascular wall [18, 28]. The Young’s modulus of the vessel wall was prescribed to be 1.2 MPa based on tensile testing performed previously on the 3D-printed material of the aortic phantoms [21]. The thickness of the wall was set to be 2.5 mm, to match the wall thickness of the 3D-printed phantoms. A Poisson’s ratio 0.5 and density 1 g/cm^3^ were used to further define the vessel wall material properties. The fluid was prescribed to have density 1.06 g/cm^3^ and viscosity 0.04 Poise to approximate the properties of blood. 3D FSI simulations were run for 10 cardiac cycles to ensure that the pressures reach periodic convergence; only the final cardiac cycle was analyzed. We report the peak pressure drop between AAo and DAo from the last cardiac cycle averaged over the cross-sectional area at the same slice locations where the catheter measurements were acquired *in vitro* (Δ*P* _FSI_).

### 2.7 Analysis

Given the limited sample size of eight datapoints, we used bootstrapping to estimate the population mean and standard deviation of the error in Δ*P* between each method and direct catheter measurements.

Agreement of Δ*P* estimates from 4D-Flow, 4D-FlowP, and FSI with catheterization measurements was evaluated using linear regression and Bland-Altman analysis. To determine the presence of systematic errors, the slope of each regression line was compared to unity (slope = 1) using a two-tailed *t*-test. Fixed bias was assessed by performing a one-sample *t*-test on the mean differences (Method *−* Cath) to determine if they significantly deviated from zero. Normality of both regression residuals and Bland-Altman differences was confirmed via Shapiro-Wilk tests (*p >* 0.05 for all methods) to justify the use of parametric statistics. All analyses were performed with a significance level of *α* = 0.05.

## 3 Results

Eight individual experiments and corresponding simulations were conducted. Tuned resistive and capacitive elements matched *in vitro* baseline pressures to clinical systolic measurements within 5 mmHg for both patients. Scan times for 4D-Flow and 4D-FlowP were comparable - 24 minutes for 4D-Flow versus 26 minutes for 4D-FlowP. A qualitative comparison of velocity fields from conventional 4D-Flow, 4D-FlowP, and FSI at the timepoint of peak velocity are shown in Figure 4.

**Fig. 4.**
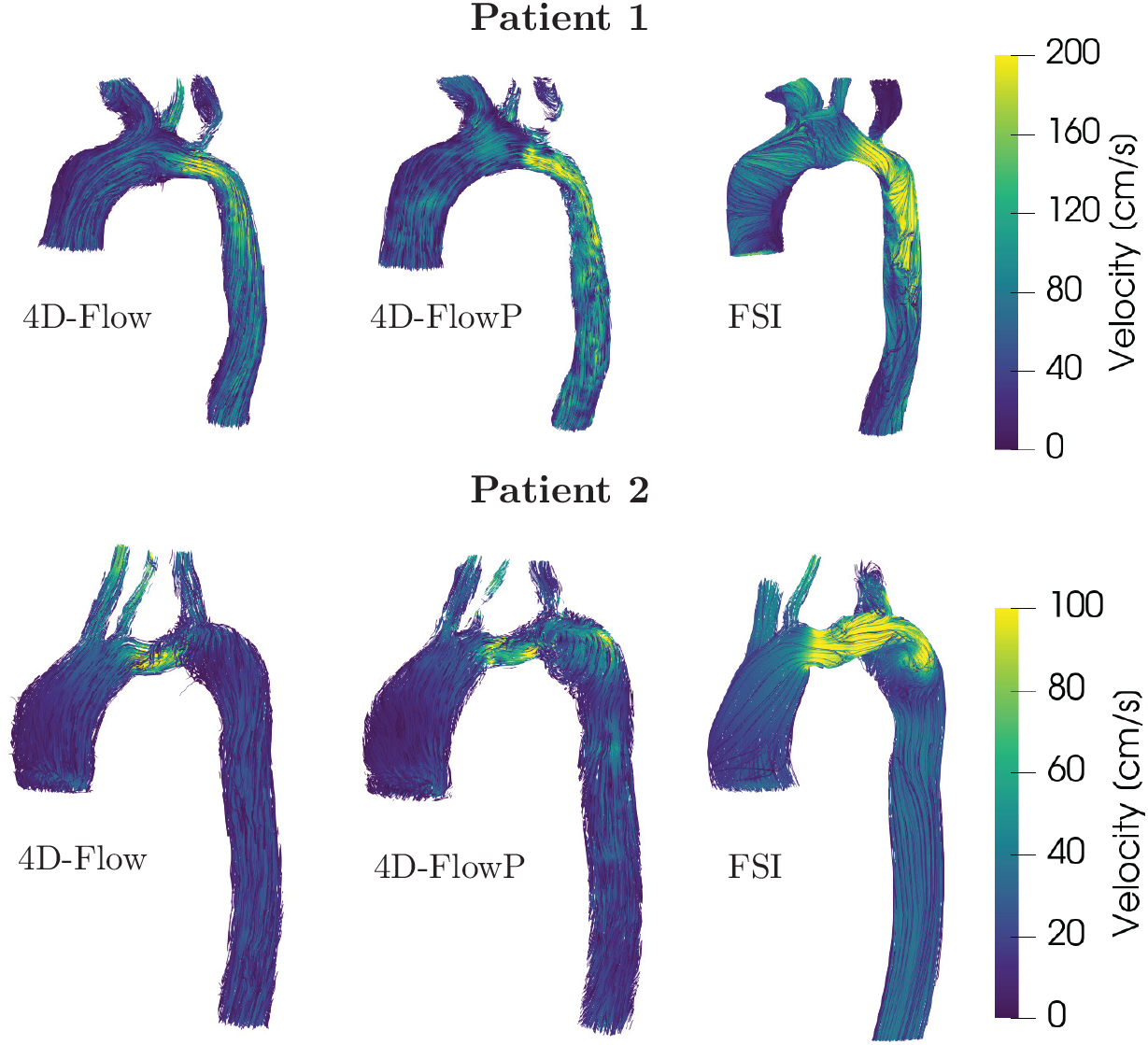
Velocity fields measured using conventional 4D-Flow (left), 4D-FlowP (middle) and FSI simulations (right) at the timepoint of peak velocity in two patients with CoA.

Qualitative comparison of the velocity fields demonstrates that 4D-FlowP effectively captures the primary flow features observed in conventional 4D-Flow MRI. 4D-FlowP velocity field reconstructions maintain spatial agreement with the reference 4D-Flow data, particularly in identifying the regions of accelerated flow and jet formation at the CoA. FSI simulations leverage higher spatial resolution to resolve more intricate secondary flow patterns. Consequently, the FSI simulations provide a more detailed look at localized velocity gradients that may be smoothed or averaged within the voxel constraints of the MRI-based methods.

The timepoint of maximum Δ*P* precedes the timepoint of peak velocity. The observed temporal offset is consistent with the Navier-Stokes momentum balance in pulsatile flow. In the accelerating phase of systole, the local acceleration term (∂**u***/*∂*t*) contributes significantly to the total pressure gradient. Consequently, the maximum pressure drop occurs during the period of greatest fluid acceleration, which inherently precedes the peak convective velocity in the phantom.

A qualitative comparison of relative pressure field maps determined from 4D-Flow, 4D-FlowP, and FSI is shown in Figure 5. In both examples, 4D-Flow underestimates the relative pressure compared to FSI simulation results. 4D-FlowP more closely matches the FSI results. We also assessed the accuracy of Δ*P*_4D-Flow_, Δ*P*_4D-FlowP_, and Δ*P*_FSI_ by comparison to Δ*P*_Cath_. The individual peak pressure drops across both patients, geometries, and flow rates are detailed in Table 3. For the whole dataset, the accuracy of each method was evaluated using linear regression plots shown in Figure 6. Conventional 4D-Flow significantly underestimated Δ*P* (*y* = 0.63*x −* 1.33, *R*^2^ = 0.75), with a slope significantly different from unity (*p* = 0.046). Conversely, 4D-FlowP (*y* = 0.95*x* + 3.74, *R*^2^ = 0.75) and FSI (*y* = 1.14*x −* 1.96, *R*^2^ = 0.82) showed no significant deviation from the ideal slope (*p* = 0.833 and *p* = 0.545, respectively). While 4D-FlowP closely tracked the gold-standard slope, its 3.74 mmHg intercept suggests a consistent overestimation at lower pressure ranges, whereas FSI provides the best overall fit.

**Fig. 5.**
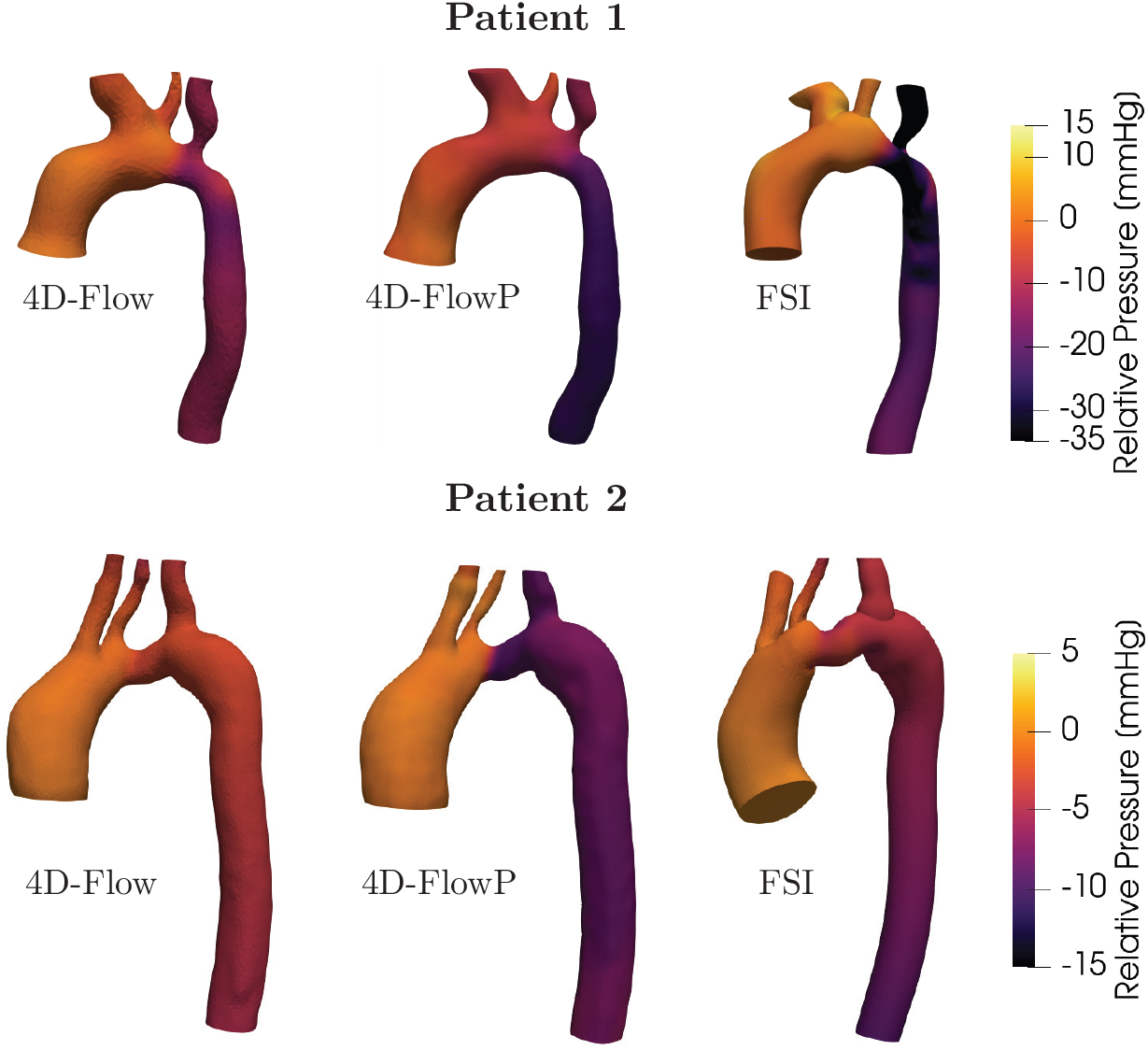
Relative pressure fields derived from conventional 4D-Flow velocity estimates (left), velocity and acceleration measurements from 4D-FlowP (middle) and FSI simulations (right)

**Fig. 6.**
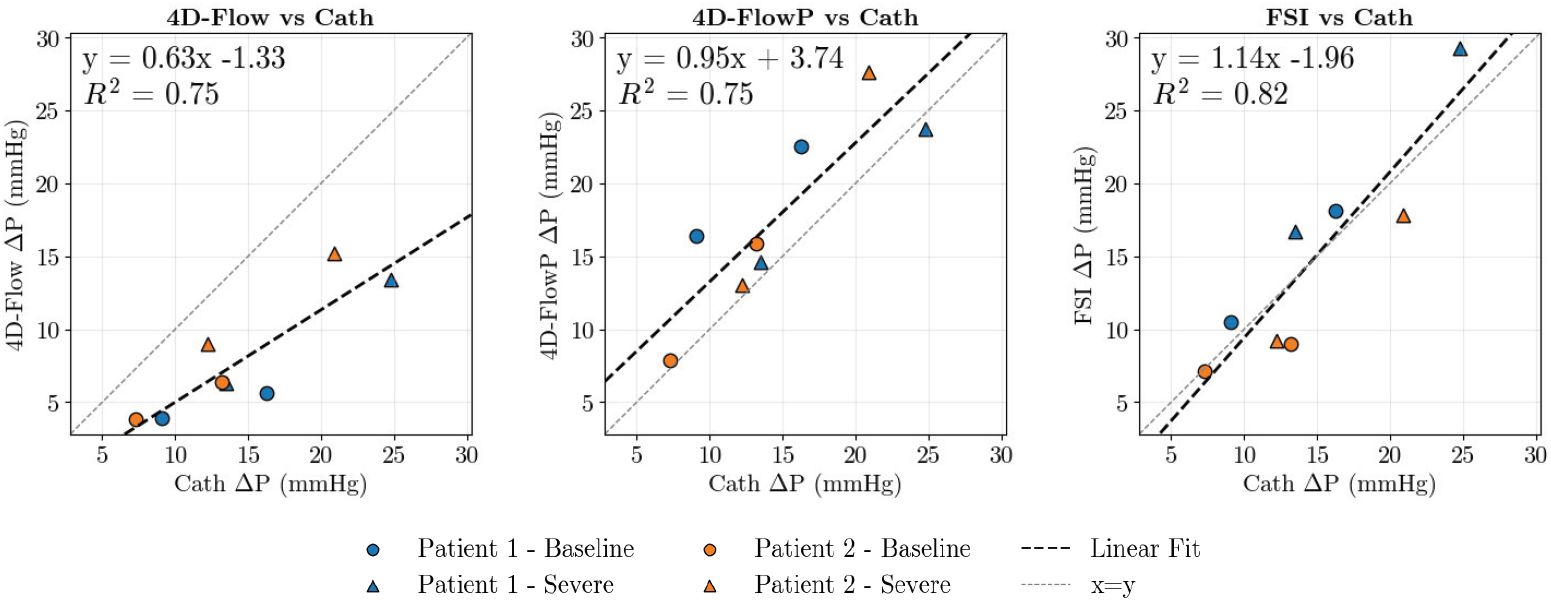
Comparison of pressure drop estimated in CoA phantoms using conventional 4D-Flow (left), 4D-FlowP (middle), and FSI simulations (right) with catheter-derived pressure measurements. Dashed line represents the linear regression.

**Fig. 7.**
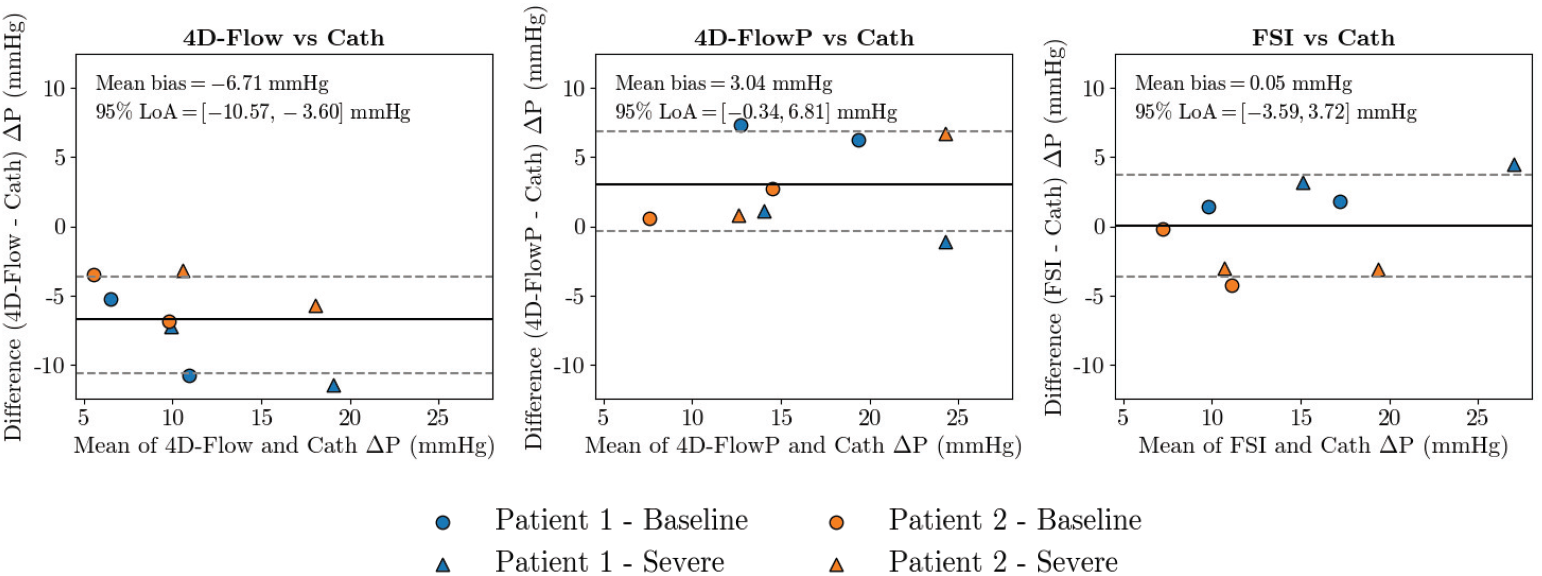
Bland-Altman plots comparing pressure gradient (Δ*P* ) estimates from conventional 4D-Flow (left), 4D-FlowP (middle) and FSI simulations (right) to catheter-derived Δ*P*_*Cath*_. The solid lines represent the mean difference between the two methods, and the paired dotted lines correspond to the 95% limits of agreement(LoA).

The agreement between Δ*P*_4D-Flow_, Δ*P*_4D-FlowP_, Δ*P*_FSI_ and ground-truth Δ*P*_Cath_ was also evaluated using Bland-Altman analysis. Δ*P*_4D-Flow_ exhibited a significant systematic underestimation, with a mean bias of -6.71 mmHg (*p <* 0.001) and 95% limits of agreement (LoA) of [-10.57, -3.60] mmHg. Δ*P*_4D-FlowP_ demonstrated a smaller but still statistically significant positive bias of 3.04 mmHg (*p* = 0.033), with a 95% LoA of [-0.34, 6.81] mmHg. FSI simulations provided the most accurate estimates of catheter-derived Δ*P*, with a near-zero bias of 0.05 mmHg that was not significantly different from zero (*p* = 0.966) and a 95% LoA of [-3.59, 3.72] mmHg.

## 4 Discussion

This study demonstrates the feasibility and accuracy of a numerically optimal, joint velocity and acceleration encoded MRI strategy (4D-FlowP) for non-invasive pressure gradient mapping in models of patients with CoA. Using patient-specific compliant phantoms under physiologically realistic flow and pressure conditions, we show that 4D-FlowP substantially improves Δ*P* estimation compared to conventional 4D-Flow MRI, with accuracy approaching that of high-fidelity FSI simulations using invasive catheter-based measurements as a reference.

Across all geometries, flow conditions, and stenosis severities studied, conventional 4D-Flow consistently underestimated Δ*P*. This finding likely highlights the sensitivity of Navier-Stokes based pressure estimation to velocity noise and temporal discretization errors, particularly in regions of high acceleration and flow separation such as post-stenotic jets. In contrast, 4D-FlowP demonstrated markedly improved agreement with catheter-derived measurements, as shown by a regression line with slope near unity, reduced bias, and narrower limits of agreement.

The improved performance of 4D-FlowP may be attributed to its simultaneous encoding of velocity and acceleration, which avoids noise amplification associated with temporal and spatial differentiation of velocity fields. By reducing reliance on numerically derived acceleration terms, 4D-FlowP mitigates one of the principal sources of error in MRI-based pressure gradient estimation. Importantly, this improvement was achieved with only a modest increase in scan time relative to 4D-Flow.

FSI simulations provided the closest overall agreement with catheter-derived pressure gradients, consistent with their ability to resolve high spatial and temporal resolution flow features. The strong correspondence between FSI-derived and catheter-measured pressure gradients supports the validity of the cardiovascular system emulator and phantom design as a realistic surrogate for patient-specific hemodynamics. 4D-FlowP matched FSI-derived pressure gradients more closely than conventional 4D-Flow across both baseline and more severe stenoses, as well as across reduced flow conditions. This suggests that the benefits of acceleration encoding are preserved across a physiologically relevant range of flow rates and pressure drops. While FSI remains a powerful validation and research tool, its computational cost, need for expert setup, and lack of immediate clinical availability limit routine use in the clinic. In this context, 4D-FlowP offers a compelling compromise between accuracy and clinical practicality. Accurate non-invasive assessment of Δ*P* remains a critical unmet need in the management of patients with CoA. Current clinical decision-making relies heavily on invasive catheterization, which exposes patients to ionizing radiation, contrast agents, procedural risks (including bleeding, infection, and vascular injury), and increased healthcare costs. Non-invasive alternatives such as Doppler echocardiography and cuff blood pressure measurements are widely used, but are known to be unreliable and often inaccurate. The results of this study suggest that 4D-FlowP has the potential to provide a more accurate, reproducible, and fully non-invasive estimate of Δ*P* that could reduce the need for diagnostic catheterization.

This study has several limitations. First, the sample size was small, with two patient-specific anatomies studied across multiple flow and stenosis conditions. While this design allowed controlled evaluation over a wide range of Δ*P*, larger *in vivo* studies are required to establish clinical robustness and usability. Second, while the phantoms were compliant and matched to physiologic aortic stiffness, they cannot fully replicate the viscoelastic and heterogeneous properties of native vascular tissue. Third, Δ*P* was computed as the peak difference between cross-sectional averages in the ascending and descending aorta, consistent with catheter measurements, but spatial pressure variations along the vessel were not evaluated in detail. Finally, although 4D-FlowP likely reduced noise-related errors, it remains sensitive to other sources of MRI uncertainty, including eddy currents, mechanical resonances, and spatiotemporal resolution limitations.

Future work will focus on *in vivo* validation of 4D-FlowP in a larger patient cohort. Establishing reliable, repeatable pressure estimates and minimizing post-processing complexity will be important prerequisites for translation to clinical practice. Beyond CoA, the ability to robustly map pressure gradients using MRI has broad applicability to other pathologies characterized by abnormal pressure gradients, such as valvular stenosis and pulmonary hypertension.

In summary, this study demonstrates that 4D-FlowP enables more accurate MRI-based pressure gradient mapping in patient-specific models of CoA than conventional 4D-Flow, with performance approaching that of invasive catheterization and FSI simulations. These findings support further clinical evaluation of 4D-FlowP as a non-invasive alternative for functional assessment of CoA severity.

## Data Availability

All data produced in the work are available online at https://doi.org/10.25740/ms528wr9373 and https://vascularmodel.org/

https://vascularmodel.org/

## Acknowledgments

This study was supported, in part, by NIH R01 HL171515 and HL183592 to DBE.

## Statements and Declarations

- Competing interests: The authors have no relevant financial or non-financial interests to disclose.
- Availability of data and materials: The simulation datasets and results generated during this study are available in the Vascular Model Repository (VMR) at vas-cularmodel.org. Model 0225 and 0232 in the VMR correspond to P-1 and P-2 in this publication. The MRI data acquired in the CoA phantoms during this study is available on the Stanford Digital Repository and can be accessed at https://doi.org/10.25740/ms528wr9373.

## Appendix

**Table 3.** *v*_*enc*_ and *a*_*enc*_ values used for in N=2 patients at two stenosis severities (Baseline and Severe) and two flow rates (*Q*_1_ and *Q*_2_)

| Patient | Geometry | Flow | $v_{enc}$<br>(cm/s) | $a_{enc}$<br>(m/s<sup>2</sup>) |
| --- | --- | --- | --- | --- |
| P-1 | Baseline | $Q_1$ | 150 | 200 |
| | | $Q_2$ | 200 | 300 |
| | Severe | $Q_1$ | 250 | 300 |
| | | $Q_2$ | 300 | 400 |
| P-2 | Baseline | $Q_1$ | 150 | 200 |
| | | $Q_2$ | 250 | 300 |
| | Severe | $Q_1$ | 250 | 300 |
| | | $Q_2$ | 300 | 400 |

